# Quality Control during the Dengue Virus Epidemic of 2024: A Multivariate Approach for Molecular Biology Diagnostics in a Multicenter Study

**DOI:** 10.64898/2026.03.18.26348458

**Authors:** Emerson Luiz Lima Araujo, Ludmila Oliveira Carvalho Sena, Jayra Juliana Paiva Alves Abrantes, Mariana Araujo Costa, Cliomar Alves dos Santos, Franciano Dias Pereira Cardoso, Josilene Felix da Rocha, Brenda Machado Moura Fernandes, Maria Gabriela Santos Silva, Erivaldo Davi dos Santos Junior, Wendel Alexandre Pinheiro de Almeida, Jean Phellipe Marques do Nascimento, Mykaella Andrade de Araujo, Hivylla Lorrana dos Santos Ferreira, Lidio Goncalves Lima Neto, Adriana Salvador, Glaucilene da Silva Costa, Jeane Maia Zeferino, Cristiane Batista Mattos, Cicileia Correia da Silva, Ediva Basilio da Silva Filho, Celina Aparecida Bertoni Lugtenburg, Daniel Ferreira de Lima Neto

## Abstract

The 2024 dengue epidemic in Brazil—the largest arboviral emergency in the country’s history—exposed critical gaps in the reliability of molecular diagnostics across its national public health laboratory network. Quality control (QC) of RT-qPCR assays performed by geographically dispersed Central Public Health Laboratories (LACENs) is essential to ensure the accuracy of epidemiological surveillance and clinical management. We conducted a multicenter QC evaluation of 3,192 complete RT-qPCR runs (19,152 datapoints) for dengue virus serotypes 1–4 (DENV1–4), Zika virus (ZIKV), and Chikungunya virus (CHIKV) across 15 LACENs over one epidemic year. An automated R-based bioinformatic pipeline applied hierarchical clustering (AGNES and DIANA), principal component analysis (PCA), linear and quadratic discriminant analysis (LDA/QDA), Shewhart and XmR control charts, process capability analysis, ANOVA, Baker’s gamma permutation testing, and PVClust bootstrap clustering to positive-control cycle threshold (CT) value datasets. Median CT values for DENV4 positive controls ranged from 26.3 to 30.5 across laboratories, representing an approximately 16-fold difference in measured RNA quantity. PCA explained 54.1%–100% of total variance on PC1 across viral targets. Baker’s gamma permutation tests confirmed significant concordance between AGNES and DIANA hierarchies across all six viral targets. LDA achieved 37.7% and QDA 49.1% cross-validated accuracy in laboratory-of-origin classification. PVClust bootstrap clustering identified DENV2+DENV4 (approximately unbiased probability, AU = 90) as the most analytically coherent serotype pair. ANOVA confirmed significant operator effects on ZIKV CT values (F = 8.799, df = 23), with regression coefficients for specific operators reaching β = +4.01 cycles—equivalent to an approximately 16-fold inferred difference in RNA quantity. Extreme outlier CT values signaled data integrity failures requiring immediate corrective action. The integrated multivariate QC framework substantially outperformed univariate Westgard-rule monitoring. Operator-specific CT deviations of up to four cycles carry direct consequences for clinical classification of borderline specimens. The automated R-based pipeline is operationally feasible in low-resource public health networks and provides a replicable model for arboviral diagnostic QC governance during epidemic emergencies.

**Author Summary:** During the 2024 dengue epidemic—the largest Brazil has ever faced—our national laboratory network processed millions of molecular diagnostic tests across 15 states. We wanted to understand whether these results, generated by different laboratories using different instruments and staff, were truly comparable to each other. We found that they were not fully interchangeable: the same standardized viral sample produced results that differed by the equivalent of a 16-fold difference in measured virus quantity, depending on which laboratory analyzed it. The main culprits were the type of machine used and the specific batch of reagents—not the geographic region or the size of the laboratory. We also found that individual technicians introduced measurable and consistent differences, some as large as four amplification cycles, which can be enough to change whether a borderline patient sample is classified as positive or negative. Using a set of advanced statistical tools routinely applied in industrial quality control, we were able to detect these patterns automatically—patterns that conventional monitoring systems would have missed. Our approach is built entirely on free, open-source software and can be adopted by any public health laboratory network in the world facing similar challenges. We believe this framework can strengthen epidemic diagnostic governance not only in Brazil but across all low– and middle-income settings.

## Introduction

Quality control (QC) is a critical component of molecular biology diagnostics, ensuring the accuracy and reproducibility of results in settings where analytical error directly affects clinical and epidemiological decision-making. In recent years, the adoption of multivariate statistical techniques has substantially expanded QC capabilities beyond the limitations of traditional univariate monitoring, enabling the detection of latent patterns, correlated sources of drift, and cross-analyte covariance shifts that per-target rule-based systems cannot capture [1].

Multivariate QC methods are particularly valuable in high-throughput molecular diagnostic platforms, where simultaneous detection of multiple targets—as in multiplex RT-qPCR assays for arboviruses—generates correlated data structures amenable to dimensionality-reduction and clustering approaches [1,2]. Multivariate approaches including Gaussian mixture models, PCA-based surveillance charts, and hierarchical clustering have demonstrated substantially improved fault-detection sensitivity relative to univariate approaches in both research and diagnostic laboratory contexts [2,3].

Hierarchical clustering algorithms—specifically agglomerative nesting (AGNES) and divisive analysis (DIANA)—represent complementary analytical strategies with distinct sensitivities to outlier structure. AGNES builds clusters bottom-up, progressively aggregating the most similar observations, making it structurally conservative and robust to localized anomalies. DIANA proceeds top-down, dividing the complete dataset by maximal internal dissimilarity, which confers greater sensitivity to early-stage deviations that have not yet disrupted the overall clustering consensus [4]. Together, they provide a methodologically triangulated view of data structure: where both algorithms agree, the identified clusters represent robust analytical groupings; where they diverge, the divergence itself encodes meaningful QC information about transitional or boundary-case observations.

Principal component analysis (PCA) reduces the dimensionality of correlated multivariate CT data onto orthogonal axes of maximal variance, enabling visualization of analytical patterns across many simultaneous variables [5,6]. In the RT-qPCR context, where CT values for multiple viral targets from the same run are expected to co-vary under common factors (reagent lot, thermocycler calibration, extraction efficiency), PCA provides a natural analytical framework for detecting systematic process shifts that manifest as coordinated multi-target CT changes.

Linear discriminant analysis (LDA) and its heteroskedastic extension, quadratic discriminant analysis (QDA), identify multivariate signatures that distinguish groups—such as laboratories or operators—enabling quantification of analytical identifiability and detection of operator-specific biases through a non-punitive, evidence-based approach [7,8]. Bootstrap-based uncertainty estimation via PVClust further allows formal probabilistic inference about the stability of hierarchical cluster structures, providing calibrated confidence measures for clustering conclusions [33].

The Westgard multi-rule system—originally developed for clinical chemistry [9]—constitutes the operational standard for per-run QC decisions in most public health laboratories. Its principal limitation in multichannel RT-qPCR settings is structural: Westgard rules monitor each analyte in isolation and are therefore blind to cross-target covariance patterns that may signal systematic drift before any individual target breaches its univariate control limits. This limitation becomes operationally consequential when multiple viral targets are detected simultaneously, as in the multiplex arbovirus assays used across the LACEN network.

The 2024 dengue epidemic in Brazil—the largest in the country’s history, with approximately 6–10 million probable cases and nearly 5,000 deaths [10]—imposed unprecedented diagnostic pressure on the national laboratory infrastructure. The simultaneous co-circulation of all four dengue serotypes alongside ZIKV and CHIKV dramatically amplified the complexity and volume of molecular diagnostic workflows [11,27]. In this context, the quality and intercomparability of RT-qPCR performance across the 15 participating LACENs acquired direct epidemiological and clinical consequences: systematic inter-laboratory CT biases affect case classification, serotype distribution estimates, and the allocation of public health resources.

In this study, we apply a comprehensive multivariate statistical framework—encompassing hierarchical clustering, PCA, LDA/QDA, ANOVA, Baker’s gamma permutation tests, PVClust bootstrap analysis, and process capability metrics—to evaluate RT-qPCR performance across the Brazilian LACEN network during the 2024 epidemic year. Our objectives were to (i) characterize the structural sources of inter-laboratory CT variability, (ii) assess whether multivariate analytical signatures of laboratory identity are detectable, (iii) quantify the contribution of operator and reagent lot effects to CT dispersion, and (iv) demonstrate the superiority of the integrated multivariate framework over conventional univariate QC approaches for epidemic diagnostic governance.

## Methods

### Study Design and Dataset

This multicenter observational QC study enrolled 15 Brazilian Central Public Health Laboratories (LACENs) participating in the national arbovirus molecular diagnostic network. The dataset comprised 3,192 complete RT-qPCR runs collected over one epidemic year (peak season through seasonal decline), representing 19,152 individual CT datapoints across six viral targets: DENV1, DENV2, DENV3, DENV4, ZIKV, and CHIKV. All RT-qPCR reactions were performed in accordance with each participating laboratory’s established protocols, using Ministry of Health-endorsed assay kits, with reagent lot numbers, run dates, operator identifiers, and thermocycler type recorded systematically for each run.

Positive controls were selected from routine RT-qPCR runs and stratified by viral target. CT values, run metadata (date, operator, reagent lot, equipment model), and quality flags were compiled into a centralized dataset for statistical analysis. Values outside pre-defined plausibility ranges (CT < 15 or CT > 38 for positive controls) were flagged for data integrity review prior to inclusion in multivariate models.

### Automated Bioinformatic Pipeline

All data preprocessing, including cleaning, transformation, and z-score normalization (scale() function), was performed using the dplyr and tidyr packages [12]. Hierarchical clustering based on multivariate CT-distance profiles was conducted using the cluster package [13], with visualization via the factoextra package [14]. AGNES (agglomerative nesting, bottom-up) and DIANA (divisive analysis, top-down) were both applied using the Manhattan distance metric and complete linkage. Agglomerative and divisive coefficients were computed as indices of clustering structure intensity. Dendrograms were compared via tanglegram visualization; concordance was quantified using the Baker’s gamma rank correlation coefficient with permutation-based significance assessment (N = 100 permutations per viral target) [4].

Bootstrap uncertainty estimation for hierarchical cluster stability was performed using the PVClust package [33], applying the approximately unbiased (AU) probability and bootstrap probability (BP) metrics across 1,000 bootstrap replicates per analysis. PCA was implemented via the factoextra package, with individual sample quality-of-representation (cos²), variable contribution, and group-level concentration ellipses visualized. LDA and QDA were performed using the MASS package [15], with leave-one-out cross-validation accuracy as the primary classification performance metric.

ANOVA (factorial design, operator as grouping variable), MANOVA, Shapiro–Wilk normality tests [17], and Levene homogeneity-of-variance tests were performed using the car package [16]. Linear regression analyses characterized operator-specific CT offsets relative to a reference operator (intercept). Quality control charts—including Shewhart X-bar, R, S, and XmR (individual measurements and moving range)—were generated using the qcc and ggQC packages [18,19]. Process capability analysis (Cpk, sigma level) was conducted for pilot laboratories SE, RN, and AC. Heatmaps of pairwise Euclidean distance matrices were generated using the ComplexHeatmap package, with DIANA-based row dendrogram construction and metadata annotation layers for laboratory, platform, reagent lot, and collection period.

## Results

### Dataset Overview and Descriptive Statistics

The compiled dataset encompassed 3,192 complete RT-qPCR runs across 15 LACENs, spanning the peak and decline of the 2024 arboviral outbreak season. Central tendency measures across all viral targets were consistent with expected performance ranges for standardized RT-qPCR assays, with narrow interquartile ranges in most laboratories indicating generally stable intra-laboratory processes. However, the presence of systematic inter-laboratory shifts in median CT values across all six viral targets indicated that local analytical stability did not translate into network-wide comparability.

The distribution of CT values for DENV4 positive controls exemplified this pattern most clearly: median CT values ranged from 26.3 (LACEN-PI) to 30.5 (LACEN-MS), representing an approximately 16-fold difference in inferred RNA quantity under the assumption of 100% amplification efficiency. Narrow intra-laboratory IQRs (2.0–3.0 cycles) confirmed that each laboratory was reproducibly measuring its own analytical level, but those levels were not interchangeable across the network.

Extreme outlier CT values warranting immediate data integrity investigation were identified in two laboratories: CT ≈ 0 in LACEN-AL and CT ≈ 18 in LACEN-RO. Both values are biologically implausible for standardized RNA positive controls and indicate severe pre-analytical or analytical failures including possible cross-contamination, instrument miscalibration, transcription errors, or RNA degradation. Their automated identification by the statistical pipeline illustrated the value of multivariate plausibility screening that would not be captured by per-run Westgard monitoring alone.

### Platform-Dependent Analytical Regimes

Stratification of CT values by thermocycler platform revealed consistent platform-dependent patterns across multiple viral targets. Laboratories using the same thermocycler system produced CT distributions that clustered closely together, while measurements generated on different instrument platforms showed systematic offsets. These offsets were reproducible across runs and persisted across all six viral targets, indicating that they arose from stable properties of the measurement system rather than from target-specific assay characteristics.

These platform-dependent analytical regimes have direct operational implications. Because instrument platform constitutes a major structural determinant of CT measurement level, diagnostic comparability across the network cannot be assumed even when identical assay kits and protocols are used. Platform identity may introduce systematic CT shifts that propagate through surveillance datasets whenever results from multiple laboratories are aggregated for epidemiological interpretation. This finding reinforces the heatmap-derived cluster structure (Cluster A: QuantStudio; Cluster B: Bio-Rad CFX; Cluster C: Applied Biosystems 7500 FAST), which consistently segregated by instrument type rather than geographic laboratory origin across all six viral targets.

### Laboratory-Level Analytical Dispersion

Stratification of CT values by individual laboratory revealed a second layer of analytical heterogeneity beyond platform-level effects. Several LACENs displayed CT distributions that were internally tight—reflected in narrow IQRs of 2.0–3.0 cycles—but systematically displaced relative to the network median. This pattern indicates that local workflows produced reproducible measurements at a measurement scale that was not interchangeable with that of other laboratories. The DENV4 target exhibited this pattern most prominently, with median CT values ranging from 26.3 (LACEN-PI) to 30.5 (LACEN-MS) (Figure 1 and 2).

**Figure 1.**
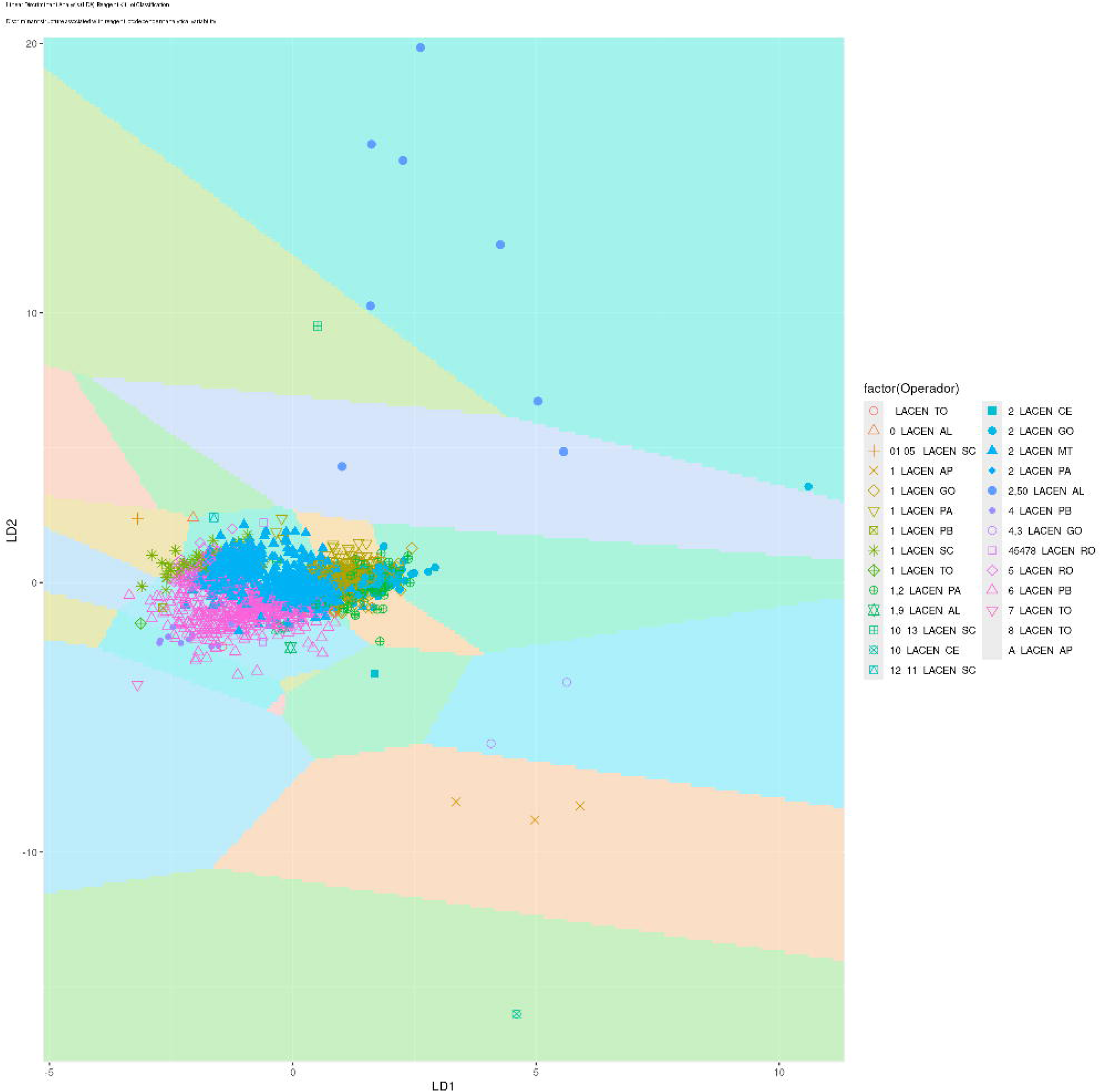
Distribution of DENV4 positive-control CT values by operator across the LACEN diagnostic network. Boxplots illustrate the dispersion of cycle threshold (CT) values obtained from standardized DENV4 positive-control reactions stratified by operator identity. Each box represents the interquartile range of CT values for a given operator, with whiskers indicating the full observed distribution and points representing outliers. The dashed horizontal line marks the network-wide median CT value. Several operators exhibit systematic CT offsets of up to four amplification cycles relative to the network median, corresponding to approximately sixteen-fold differences in inferred RNA quantity under ideal amplification efficiency assumptions. These results demonstrate that operator-specific procedural variability constitutes a measurable contributor to network-level analytical heterogeneity.

**Figure 2.**
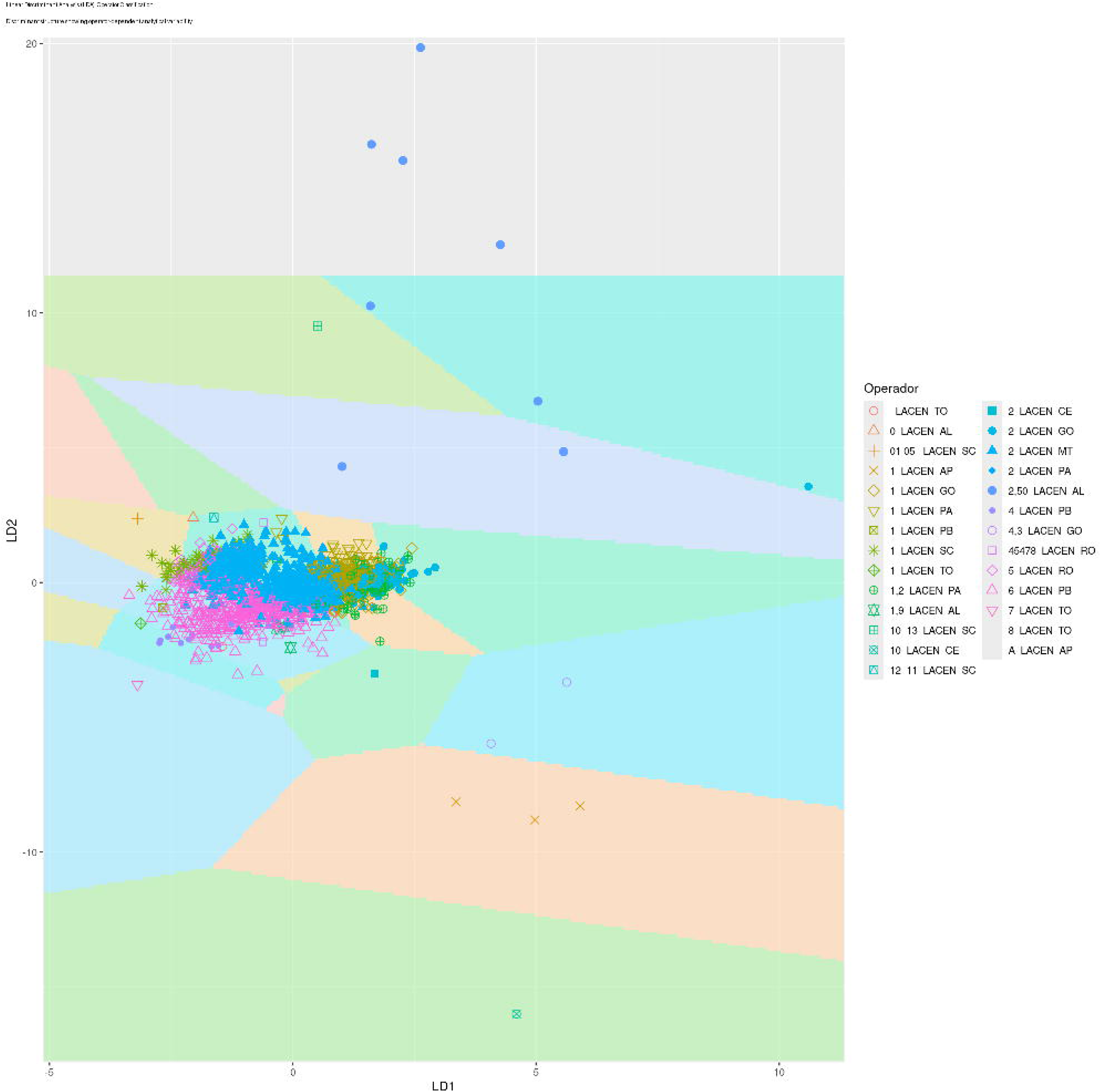
Influence of thermocycler platform on RT-qPCR positive-control CT measurements. Distribution of DENV4 positive-control CT values stratified by thermocycler model used across the participating LACEN laboratories. Distinct CT distribution regimes are evident between instrument platforms, indicating that optical detection systems, amplification kinetics, and calibration procedures contribute significantly to analytical variability within the diagnostic network. Despite standardized protocols and reagents, systematic platform-dependent CT offsets are observed, emphasizing the importance of instrument harmonization in distributed molecular diagnostic systems.

These systematic laboratory-level offsets likely reflect stable combinations of instrument calibration state, reagent lot configuration, and local workflow practices—factors that can remain constant within a laboratory over extended periods and generate analytically reproducible yet non-comparable results. The process capability analysis for pilot laboratories SE, RN, and AC confirmed this heterogeneity: laboratories with similar median CT values differed substantially in their Cpk values and temporal stability profiles, demonstrating that measurement centering and measurement precision represent independent dimensions of analytical performance requiring distinct governance strategies.

### Hierarchical Clustering and Tanglegram Analysis

AGNES and DIANA were applied to scaled CT distance matrices for each viral target independently. Agglomerative coefficients approached 1.0 across all targets, indicating highly defined clustering structure. The DIANA divisive coefficient confirmed the complementary perspective: clear internal partitioning consistent with the AGNES-derived groupings.

Tanglegram comparison of AGNES and DIANA dendrograms revealed high concordance at the highest hierarchical levels across DENV1–4, ZIKV, and CHIKV, with divergences concentrated at deeper nodes reflecting method-specific sensitivities. The dominant first-level splits consistently reflected PCR platform origin—QuantStudio vs. CFX Bio-Rad vs. Applied Biosystems 7500 FAST—confirming thermocycler type and reagent batch as the principal structural drivers of CT-distance organization (Figure 3).

**Figure 3.**
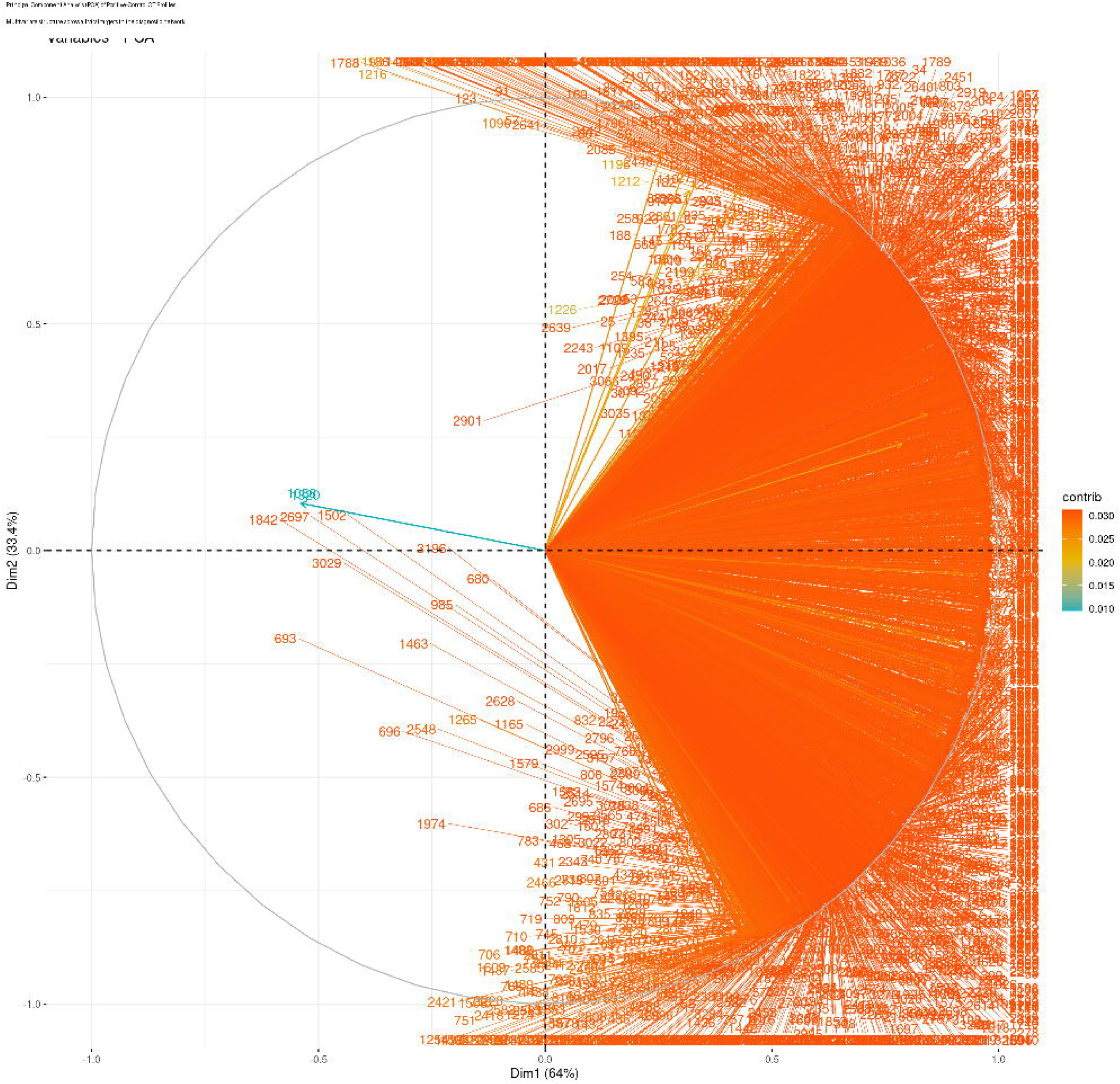
Tanglegram comparison of AGNES and DIANA hierarchical clustering structures for DENV2 positive-control CT profiles. The tanglegram visually compares dendrograms generated using the AGNES (agglomerative nesting) and DIANA (divisive analysis) hierarchical clustering algorithms applied to the same CT distance matrix. Colored lines connect identical observations across the two trees. Although extensive line crossing is visible, this pattern reflects differences in internal leaf ordering rather than disagreement about cluster membership. Baker’s gamma permutation tests confirmed statistically significant concordance between the two hierarchical structures across all viral targets, demonstrating that the analytical regimes identified by clustering represent genuine structural features of the diagnostic measurement system rather than artifacts of a specific algorithm.

A methodologically important contrast emerged in the DENV-2 analysis: DIANA early-excised an anomalous reagent lot in its first divisive step, treating it as maximally dissimilar from the core ensemble, whereas AGNES progressively incorporated the same lot into a higher-order cluster, effectively concealing its deviant character until later in the agglomeration hierarchy. This asymmetry demonstrates that divisive clustering provides earlier and more sensitive detection of emerging batch-specific deviations—a property with direct operational value for lot-qualification decisions prior to large-scale deployment of a suspect reagent batch [4,32].

The pattern of extensive visual line-crossing in tanglegrams—often interpreted as indicator of disagreement—here reflects a different phenomenon: the two algorithms identify the same global cluster structure but differ in the internal ordering of leaves within clusters. This distinction is critical for interpretation: leaf-ordering divergence is an expected consequence of the opposed directionalities of the two algorithms (bottom-up vs. top-down) and does not represent structural disagreement about cluster membership.

### Baker’s Gamma Permutation Tests

Baker’s gamma rank correlation was computed between AGNES and DIANA dendrogram pairs for all six viral targets, with significance assessed via permutation testing (N = 100) under the null hypothesis of random dendrogram concordance.

**Table 1.** Baker’s gamma permutation test results comparing AGNES and DIANA hierarchies for each viral target. All p-values equal zero under 100 permutations, indicating that AGNES–DIANA concordance significantly exceeds random expectation across all targets.

| Viral Target | Bandwidth ($H_0$) | Baker's $\gamma$ | p-value |
| --- | --- | --- | --- |
| ZIKV | 0.00468 | $\geq 0.90$ | 0 |
| CHIKV | 0.0009213 | $\geq 0.90$ | 0 |
| DENV1 | 0.006625 | $\geq 0.90$ | 0 |
| DENV2 | 0.005023 | $\geq 0.90$ | 0 |
| DENV3 | 0.001801 | $\geq 0.90$ | 0 |
| DENV4 | 0.004988 | $\geq 0.90$ | 0 |

Permutation p-values of zero (across N = 100 simulations) for all six viral targets confirmed that the AGNES and DIANA hierarchical structures are significantly more concordant than expected by chance. This result has a dual interpretation: it validates that the identified cluster structure is real rather than algorithmic artifact, and it establishes that platform– and lot-driven analytical groupings are robust across fundamentally different clustering paradigms.

### PVClust Bootstrap Analysis

Bootstrap clustering analysis (1,000 replicates) of CT profiles across all six viral targets simultaneously produced a consensus dendrogram with AU and BP confidence values for each node. The most stable node paired DENV2 and DENV4 (AU = 90, BP = 72), indicating that these two serotypes produce the most mutually similar positive-control CT profiles across the network. DENV1 and DENV3 formed the second-most-stable cluster (AU = 56, BP = 33), with ZIKV and CHIKV grouping at intermediate confidence (AU = 69 and 47, respectively).

The DENV2+DENV4 pairing (AU = 90) meets the conventional ≥0.85 AU threshold for reliable bootstrap support, reflecting genuine structural similarity in the analytical performance profiles of these two serotype assays across the network. The DENV1+DENV3 pairing (AU = 56) falls below this threshold and should be interpreted cautiously, suggesting residual uncertainty about the stability of this grouping that additional replications or expanded metadata covariates might resolve.

### Principal Component Analysis

PCA of scaled CT distance matrices across reagent lot groups for each viral target revealed that PC1 accounted for 54.1% (DENV4) to 100% (DENV3) of total variance, confirming the dominance of a single latent dimension of overall assay dispersion in all analyses (Figure 4).

**Figure 4.**
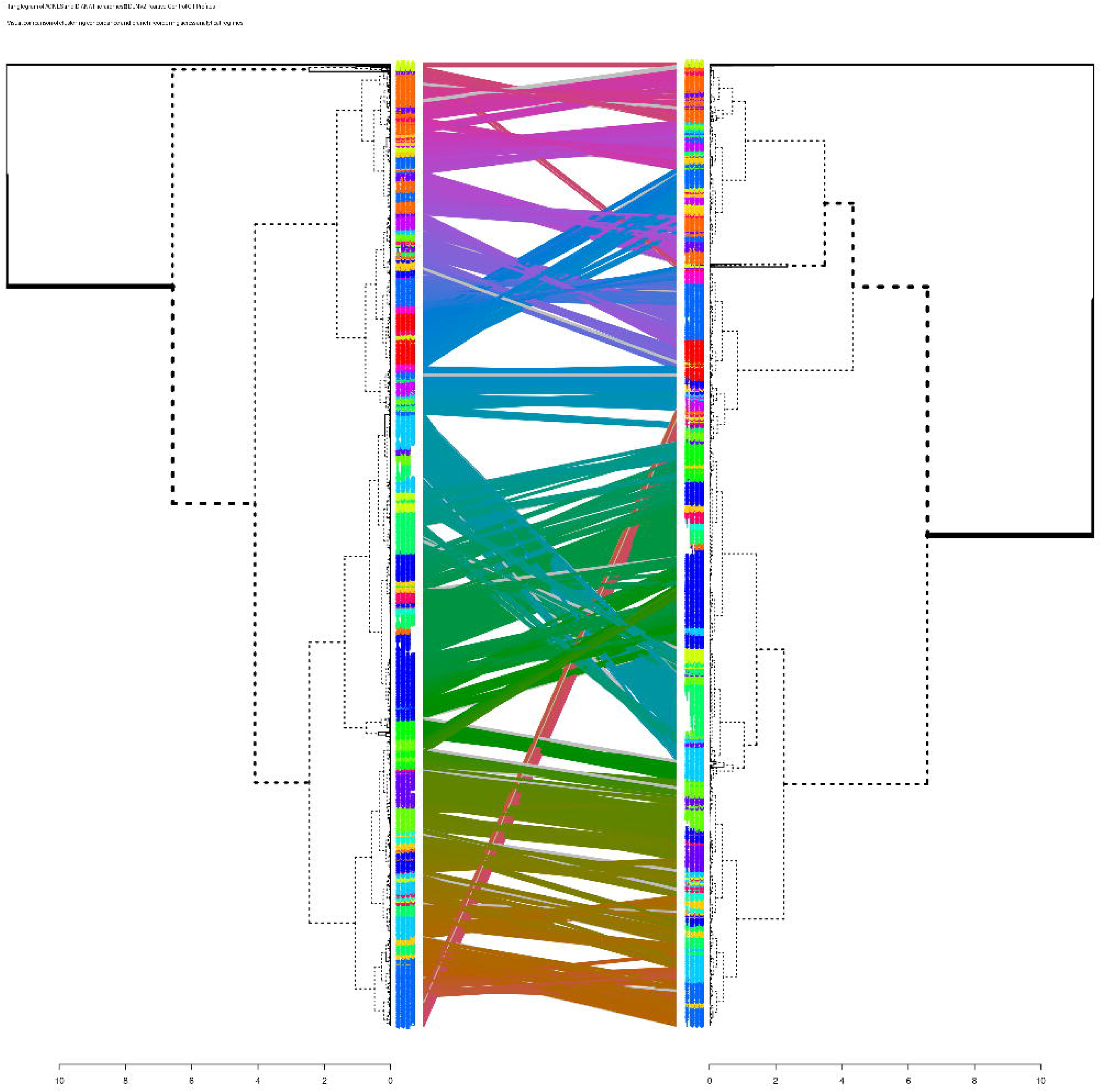
Principal component analysis of multivariate CT profiles across the diagnostic network. Projection of scaled CT distance matrices into principal component space summarizing multivariate relationships among RT-qPCR runs across all viral targets. Each point represents an individual RT-qPCR run. PC1 accounts for the majority of total variance in the dataset (ranging from 54.1% to 100% depending on viral target), indicating the presence of a dominant latent dimension representing overall assay dispersion. The concentration of most observations within a narrow region of PCA space reflects general analytical consistency, whereas peripheral points represent outlier runs potentially associated with reagent transitions, instrument recalibration events, or procedural deviations.

**Table 2.** PCA variance explained by PC1 and PC2 for each viral target. The combined two-component solution captures ≥96% of total variance across all targets.

| Viral Target | PC1 Variance (%) | PC2 Variance (%) | PC1 + PC2 (%) |
| --- | --- | --- | --- |
| ZIKV | 62.6 | 34.1 | 96.7 |
| CHIKV | 68.2 | 28.8 | 97.0 |
| DENV1 | 71.8 | 25.5 | 97.3 |
| DENV2 | 62.1 | 34.3 | 96.4 |
| DENV3 | 72.8 | 24.7 | 97.5 |
| DENV4 | 62.6 | 34.1 | 96.7 |

DENV3’s exceptional uniformity (PC1 = 100%, negligible PC2) establishes it as a reference anchor for assessing reagent batch performance. DENV4’s more distributed variance structure (PC1 = 54.1%, PC2 = 34.1%) indicates a more complex multi-factor variability landscape. For ZIKV and CHIKV, the substantial PC2 contribution reflected systematic directional CT biases—consistent positive or negative shifts across replicates—attributable to primer-probe thermodynamic changes or operator-dependent pipetting patterns. A single anomalous DENV1 lot was identified at extreme PC1/PC2 coordinates as a priority audit target.

PCA stratified by operator revealed a characteristic arc-shaped distribution for ZIKV, CHIKV, DENV1, and DENV3, with the majority of operators concentrated in a central dense region and a small number of outlier operators occupying extreme positions along PC1. PCAs stratified by reagent lot for DENV1–DENV4 showed overlapping confidence ellipses with displaced centroids between lots, confirming that lot transitions introduce measurable CT shifts even within individual laboratories. Recurrent outlier run IDs (1183, 1200, 1202, 1204, 1213, 1214, 1259, 1264, 1285) appeared as isolated points across multiple independent viral-target PCAs, warranting specific root-cause investigation.

### Linear Discriminant Analysis

LDA and QDA classification of RT-qPCR runs by laboratory of origin yielded cross-validated accuracies of 37.7% and 49.1%, respectively. Both values significantly exceed random expectation for a 15-class problem (6.7% chance accuracy), confirming that CT profiles encode detectable laboratory-specific signatures) (Figure 5).

**Figure 5.**
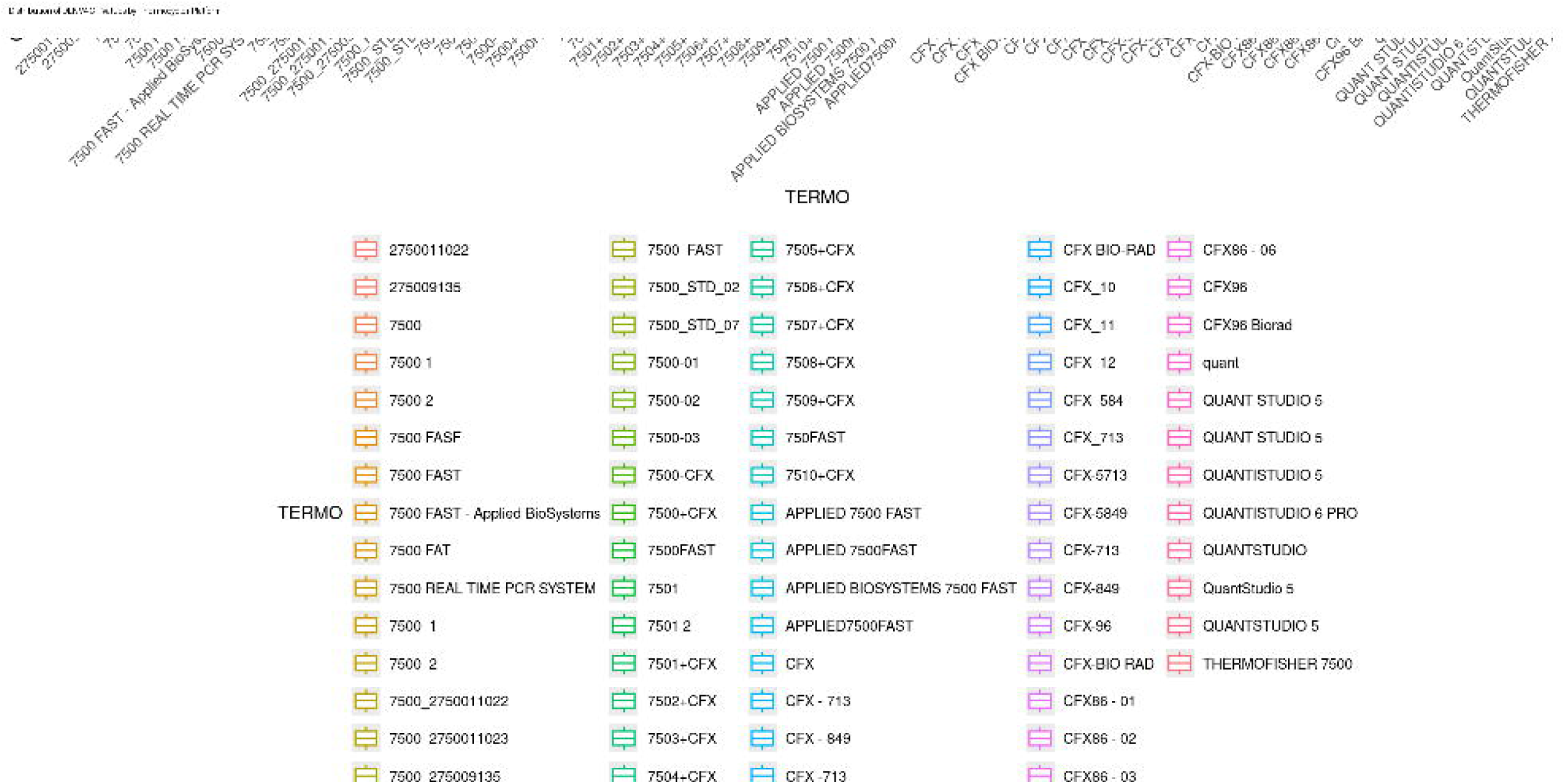
Linear discriminant analysis of reagent kit lot identity. LDA projection illustrating the discriminant structure associated with reagent lot groups used in RT-qPCR assays across the LACEN network. Background color regions represent predicted classification zones for each reagent lot category, while points correspond to individual RT-qPCR runs. Partial separation between groups indicates that reagent chemistry introduces systematic analytical signatures detectable in CT profiles. These results confirm that reagent lot transitions represent an important source of measurement variability within distributed diagnostic systems.

The superior QDA performance (Δaccuracy = +11.4 percentage points) indicates that inter-laboratory variability involves heterogeneous covariance structures—each laboratory’s CT values for different viral targets co-vary distinctively—rather than merely mean-level shifts across a shared covariance framework. This heteroskedastic pattern characterizes multicenter molecular diagnostic environments where equipment, protocols, and environmental conditions create distinctive analytical fingerprints [37,38].

In LD1–LD2 space, the majority of operator-laboratory combinations clustered tightly around the origin, reflecting overlapping multivariate CT profiles. A small number of combinations—notably “2_LACEN-GO” and “1_LACEN-AP”—occupied distinctly remote positions, indicating systematically reproducible deviations consistent with operator-specific bias or protocol inconsistency. An extreme outlier at LD2 ≈ +20 for “2_LACEN-MT” almost certainly represents a data integrity failure rather than biological variation. CT_ZIKA and CT_DEN2 contributed most strongly to discriminant loadings, designating these targets as the most sensitive markers of laboratory-specific procedural biases (Figure 6).

**Figure 6.**
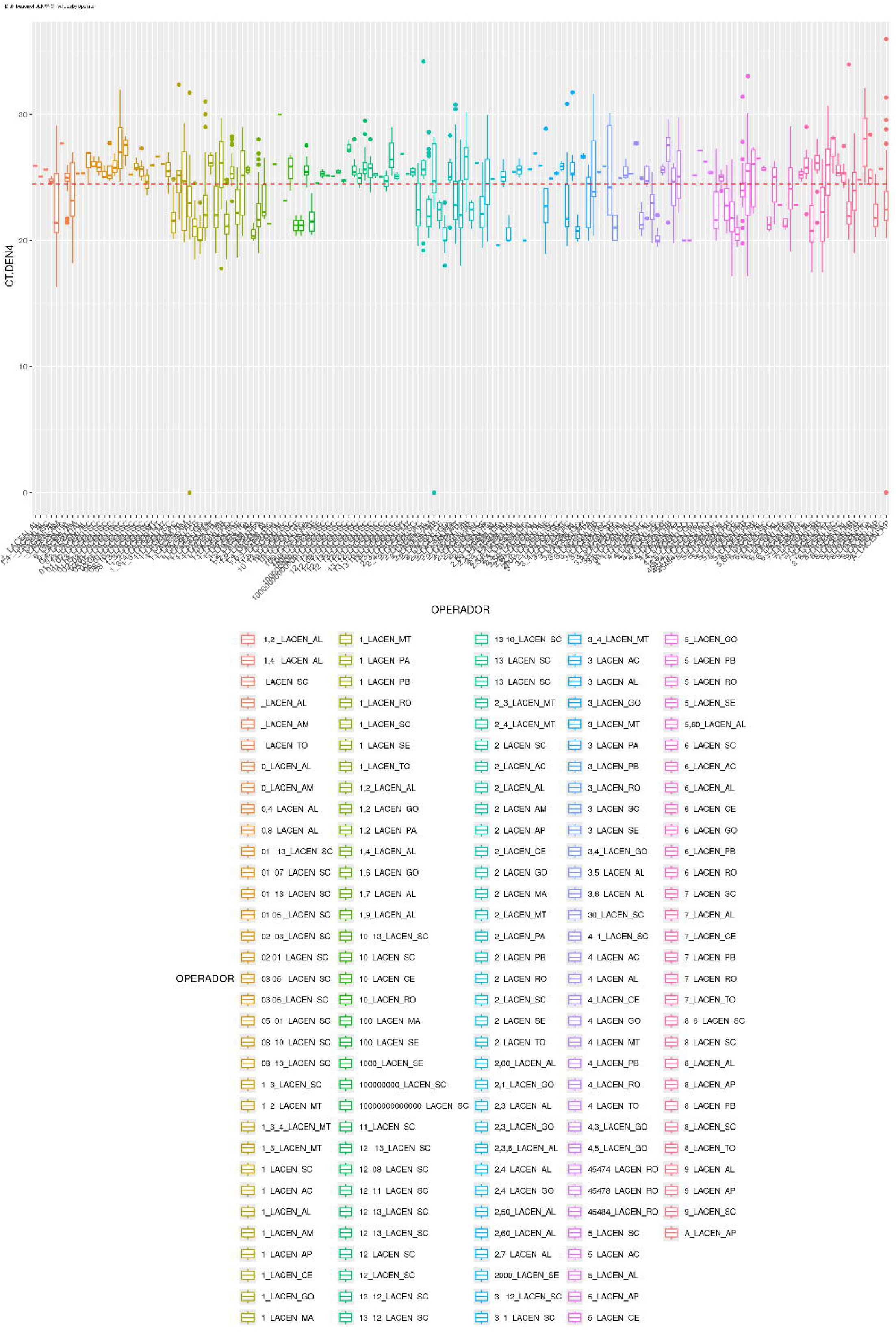
Linear discriminant analysis of operator-dependent analytical variability. LDA projection illustrating classification of RT-qPCR runs according to operator identity. Each symbol corresponds to an individual run colored by operator group. Although clusters partially overlap due to shared protocols and reagents, the discriminant axes reveal measurable operator-specific analytical signatures in CT measurements. The presence of distinct classification regions confirms that human procedural variability contributes significantly to the analytical structure of the diagnostic network.

### ANOVA, Regression, and Operator Effects

ANOVA with operator as grouping variable confirmed a significant operator effect on ZIKV CT values (F = 8.799, df = 23, p ≈ 0), accounting for a mean square of 93.97 against residual mean square of 10.68. Levene’s test confirmed heterogeneous variances across operators (statistic = 3.680, p ≈ 0, df = 23), and the Shapiro–Wilk test detected significant non-normality of residuals (W = 0.850, p ≈ 0)—violations that motivated recommendation of complementary non-parametric testing.

Linear regression of CT_ZIKA on operator identity (with a reference operator as intercept = 22.017 cycles) identified multiple operators with individually significant positive CT offsets:

**Table 3.**
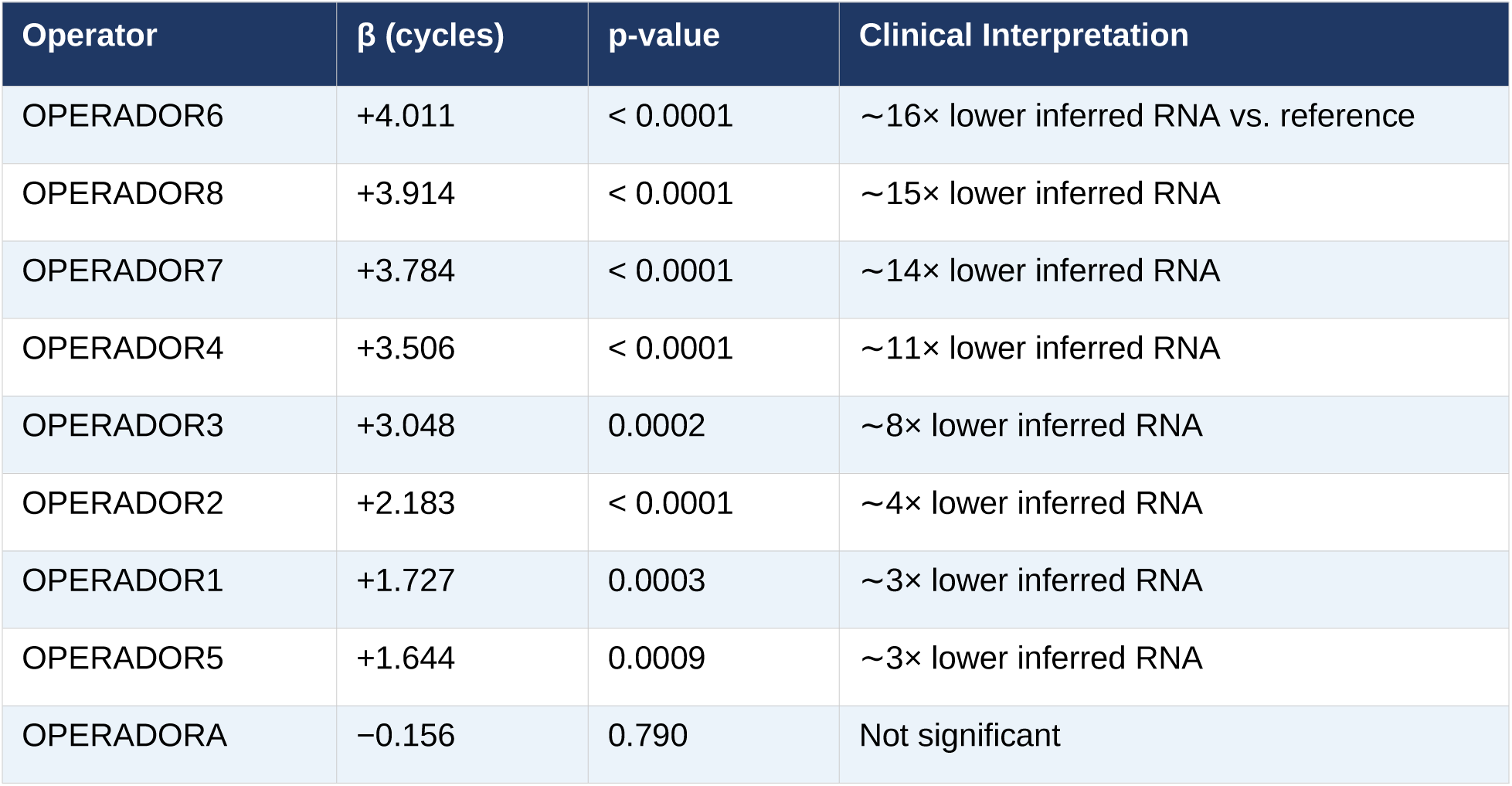
Significant operator-specific regression coefficients for CT_ZIKA. β values represent cycles above the reference operator intercept (22.017 cycles). Clinical interpretation assumes 100% amplification efficiency (each ΔCT = 1 corresponds to 2× difference in RNA quantity).

| Operator | $\beta$ (cycles) | p-value | Clinical Interpretation |
| --- | --- | --- | --- |
| OPERADOR6 | +4.011 | < 0.0001 | ~16× lower inferred RNA vs. reference |
| OPERADOR8 | +3.914 | < 0.0001 | ~15× lower inferred RNA |
| OPERADOR7 | +3.784 | < 0.0001 | ~14× lower inferred RNA |
| OPERADOR4 | +3.506 | < 0.0001 | ~11× lower inferred RNA |
| OPERADOR3 | +3.048 | 0.0002 | ~8× lower inferred RNA |
| OPERADOR2 | +2.183 | < 0.0001 | ~4× lower inferred RNA |
| OPERADOR1 | +1.727 | 0.0003 | ~3× lower inferred RNA |
| OPERADOR5 | +1.644 | 0.0009 | ~3× lower inferred RNA |
| OPERADORA | -0.156 | 0.790 | Not significant |

These findings confirm that operator identity is a significant determinant of absolute CT measurements in the LACEN network. The clinical significance of these offsets is substantial: a β of +4.0 cycles implies that an operator-specific CT of 30 for a specimen with true threshold-level viral load could be classified as positive by a reference operator (CT = 26), illustrating a direct pathway from operator-level variability to diagnostic misclassification.

### Post-Hoc Operator Comparisons

Pairwise post-hoc comparisons using Tukey’s Honest Significant Difference (HSD) test were performed to identify specific operator pairs with statistically distinguishable CT profiles. A substantial number of operator pairs displayed measurable CT differences, although many comparisons produced wide confidence intervals attributable to unequal sample sizes across operators. This pattern indicates that operator-dependent analytical variation is distributed across the network rather than concentrated within a single laboratory or instrument group.

Importantly, these pairwise differences do not necessarily indicate procedural non-compliance. They more likely reflect systematic micro-variations in pipetting technique, reagent preparation, sample handling, and extraction procedure that propagate through amplification cycles in a consistent, operator-specific manner. Even under identical written protocols, such micro-variations generate detectable and reproducible CT signatures when analyzed at the scale of the full network dataset. This interpretation is consistent with the regression findings: operator-specific β offsets were stable and individually significant, indicating structured rather than random sources of variability.

Note that the parametric Tukey HSD test assumes equal variances; given the confirmed heteroskedasticity across operators (Levene statistic = 3.680, p ≈ 0), Games–Howell post-hoc testing is recommended as a variance-robust alternative for future pipeline iterations. Kruskal–Wallis and Welch ANOVA should similarly be incorporated to validate the ANOVA conclusions under the non-normality conditions documented by the Shapiro–Wilk test (W = 0.850, p ≈ 0).

### Process Capability and Control Charts

Process capability analysis (Cpk, sigma level) for pilot laboratories SE, RN, and AC quantified alignment of CT distributions with pre-specified specification limits. XmR and Shewhart control charts revealed both stable and drift-prone patterns across the epidemic year, with temporal shifts attributable to reagent lot changes, equipment recalibration events, or seasonal environmental factors.

A critical finding from this analysis was that laboratories with comparable overall CT medians differed substantially in Cpk values, confirming that mean CT is an insufficient sole QC metric. Some laboratories displayed stable but systematically biased CT distributions (consistent with a calibration offset), while others showed accurate medians but elevated within-run variability. These structural differences require distinct corrective strategies: offset bias is best addressed through inter-laboratory CT harmonization protocols, while elevated variability requires process stabilization through reagent standardization or operator training.

### Heatmap Analysis

ComplexHeatmap visualization of pairwise Euclidean distance matrices for all six viral targets, annotated by laboratory origin, PCR platform, reagent lot, and collection period, revealed three principal structural groupings consistent across targets. Cluster A comprised predominantly QuantStudio-derived controls (LACEN-SC and MT; intra-cluster distances ≤ 1.5); Cluster B included Bio-Rad CFX-processed controls (LACEN-AL, PA, GO; distances 1.5–2.5); and Cluster C consisted of controls processed on older Applied Biosystems 7500 FAST instruments (LACEN-SE, SC; distances 2.5–4.0).

The DENV3 heatmap was notable for a distinct sub-cluster exhibiting elevated distances (red/orange tones in the visualization scale) relative to the broader network ensemble—consistent with either a problematic reagent lot or a divergent protocol that had not been flagged through univariate Westgard monitoring. A subset of five to seven ZIKV outlier samples with pairwise distances ≥ 4.5, concentrated in early QuantStudio runs, was identified as requiring re-testing to exclude RNA degradation, mislabeling, or contamination. The temporal concentration of these outliers in a discrete subperiod suggested a batch-specific reagent transition rather than ongoing operational failure.

### Implications for Diagnostic Surveillance

The results presented above demonstrate that the LACEN network maintained operational stability and sustained large-scale diagnostic throughput throughout the 2024 epidemic, while simultaneously exhibiting measurable analytical heterogeneity at the network level. For clinical diagnostics performed within individual laboratories, such within-laboratory variability may have limited practical consequences as long as local control limits are observed. However, for epidemiological surveillance systems that aggregate RT-qPCR results across laboratories, the systematic CT shifts documented here may influence quantitative interpretations of viral circulation patterns and case classification.

In particular, diagnostic thresholds used to classify specimens as positive, indeterminate, or negative may be differentially applied depending on which laboratory performs the assay, given the platform– and operator-dependent CT offsets identified in this study. Specimens with CT values near classification boundaries are especially vulnerable to such effects. These findings underscore that network-level analytical harmonization is as important as individual laboratory quality control for surveillance systems that derive epidemiological inferences from aggregated molecular diagnostic data. The multivariate statistical framework implemented here provides a practical, open-source-based tool for continuously monitoring analytical regimes across the network, enabling early detection of emerging biases before they propagate into surveillance estimates.

### The LACEN Network as a Diagnostic Measurement Ecosystem

The integrated results of this study support a conceptual reframing of distributed diagnostic networks as measurement ecosystems in which analytical results emerge from interactions among multiple co-occurring technical components. Instruments, reagent chemistry, operator execution, software algorithms, and environmental conditions collectively shape the CT values produced by RT-qPCR assays. These components do not operate independently: platform-specific detection algorithms interact with master mix fluorescence characteristics, operator pipetting precision interacts with reagent lot stability, and thermocycler calibration interacts with probe kinetics at each amplification cycle. The patterns identified through hierarchical clustering, PCA, and discriminant analysis in this study reflect the aggregate output of these interactions across fifteen distinct laboratory environments operating simultaneously within a single epidemic surveillance network.

Within this ecosystem, analytical variability was not uniformly distributed: platform identity and reagent lot identity consistently emerged as the dominant structural drivers, with operator effects constituting a significant secondary layer. Geographic proximity or institutional affiliation did not predict analytical similarity. This pattern confirms that the primary governance challenge for distributed molecular diagnostic networks is not one of institutional management but of technical-operational harmonization: standardizing instrument platforms, reagent qualification procedures, and operator training protocols constitutes the most analytically impactful intervention available to network administrators. Epidemiological interpretation of nationally aggregated RT-qPCR data must account for this underlying technical heterogeneity, particularly when deriving quantitative inferences such as viral load distributions, threshold-based case prevalence, or serotype-specific incidence estimates.

## Discussion

The unprecedented dengue epidemic that affected Brazil in 2024 placed extraordinary pressure on the national arboviral diagnostic infrastructure. Under such conditions, diagnostic systems transition from routine clinical workflows to large-scale epidemiological measurement platforms. This transition fundamentally changes the role of molecular diagnostics: RT-qPCR assays no longer function solely as clinical decision tools but also as quantitative sensors of epidemic dynamics. Ensuring the analytical comparability of measurements across geographically distributed laboratories therefore becomes critical for accurate epidemiological surveillance. The present study provides a comprehensive multivariate assessment of molecular diagnostic performance across the Brazilian LACEN laboratory network during this epidemic, revealing an important structural property of distributed molecular diagnostic networks: local analytical stability does not necessarily imply network-wide comparability.

The distinction between reproducibility and comparability represents one of the most significant conceptual outcomes of this work. Most laboratories in the network displayed relatively stable intralaboratory Ct distributions, suggesting that internal workflows were functioning consistently within their local environments. However, when these distributions were compared across the national network, systematic differences became evident. Because RT-qPCR amplification follows an exponential process, small cycle differences correspond to substantial changes in inferred nucleic acid quantity [29]. Under ideal amplification efficiency assumptions, a difference of four cycles corresponds to roughly a sixteen-fold difference in RNA concentration. Such variation illustrates that results produced in different laboratories may not be directly interchangeable even when each laboratory individually satisfies internal quality criteria.

This phenomenon is not unique to arbovirus diagnostics. Interlaboratory studies in molecular testing have repeatedly demonstrated that identical assays can yield different quantitative outputs when performed across different instruments, reagent formulations and laboratory environments. Cross-platform comparisons of qPCR assays have shown that thermocycler platform and master mix composition can produce significant differences in Ct values even when identical primer–probe systems are used [52]. Similarly, methodological studies of RT-qPCR variability emphasize that amplification efficiency, background correction and normalization procedures can introduce substantial analytical variation if not carefully standardized [53]. Taken together, these findings underscore the inherent sensitivity of RT-qPCR measurements to technical context and validate the network-level patterns we observed in the LACEN dataset.

In distributed diagnostic networks, such sensitivity creates a fundamental challenge. Each laboratory operates within a unique combination of hardware, reagents, workflow design, and personnel expertise---what may be described as a complex measurement ecology. Instruments, reagent chemistry, operator execution, software algorithms, and environmental conditions collectively shape the CT values produced by RT-qPCR assays. Even when standardized protocols are used, small differences in thermocycler calibration, fluorescence detection algorithms, amplification efficiencies, or reagent chemistry can introduce measurable shifts in CT values. The coexistence of heterogeneous configurations within a single surveillance network can therefore generate latent variability that only becomes apparent when data are aggregated at the national scale---a pattern consistent with what has been termed the reproducibility–comparability gap in multicenter molecular diagnostics. The results presented here demonstrate that this phenomenon occurred during the 2024 dengue epidemic: laboratories were internally consistent yet collectively heterogeneous.

The multivariate analyses performed in this study provide insight into the underlying structure of this heterogeneity. Hierarchical clustering consistently revealed patterns associated with thermocycler platform and reagent lot rather than geographic location. This observation suggests that analytical variability in the LACEN network arises primarily from technical-operational factors rather than from regional or institutional characteristics. Previous studies have reported similar findings in molecular diagnostics: variability in RT-qPCR results has been attributed to differences in reagent chemistry, fluorescence detection systems, amplification efficiency and calibration algorithms, all of which can influence CT determination independently of geographic or institutional variables [53].

Another important contributor to analytical variability appears to be operator-dependent execution. Although modern RT-qPCR workflows aim to minimize manual variation through standardized protocols and automation, the technique still involves numerous steps that depend on human execution. Pipetting accuracy, reagent handling, plate preparation and nucleic acid extraction procedures can all introduce subtle variations that propagate through amplification cycles. Studies of RT-qPCR variability in high-throughput surveillance contexts have demonstrated that operator effects, including differential pipetting precision and extraction technique, represent a measurable component of total variability even under otherwise controlled conditions [55]. Our ANOVA results (F = 8.799, p ≈ 0, df = 23) and regression coefficients reaching β = +4.0 cycles confirm that these operator effects have clinical-scale consequences in the LACEN network.

The epidemiological consequences of such variation are nontrivial. Diagnostic classification in many surveillance systems relies on CT thresholds that separate positive from negative or indeterminate results. If systematic operator– or platform-specific biases shift CT measurements by several cycles, identical clinical samples may be classified differently depending on where they are tested. For specimens with CT values near diagnostic thresholds, such biases could lead to misclassification, potentially affecting case counts, serotype distribution estimates, and outbreak trajectory analyses. A β of +4.0 cycles, as observed for specific operators in LACEN-associated ZIKV testing, implies that a specimen with a true threshold-level viral load (CT = 26 for a reference operator) would yield CT = 30 under that operator’s execution—potentially shifting classification from positive to indeterminate or negative depending on the cutoff applied. This represents a direct pathway from operator-level analytical variability to diagnostic misclassification with population-level consequences.

The discriminant analyses conducted here further suggest that laboratory signatures are not limited to simple shifts in mean CT values. Linear discriminant analysis achieved a cross-validated classification accuracy of 37.7%, while quadratic discriminant analysis improved this to 49.1%—both values substantially exceeding the random expectation for a fifteen-class problem (6.7%). The superior performance of QDA (Δ = +11.4 percentage points) supports a key interpretation: laboratories appear to differ not only in their mean CT levels but in their covariance structures, indicating that CT values across viral targets vary together in different ways depending on the analytical environment. QDA allows each group to possess its own covariance matrix, making it better suited to detect heteroscedastic laboratory signatures. In practical terms, this implies that multivariate quality surveillance must account for second-order statistical structure, not merely mean offsets [37,38].

This observation highlights an important limitation of traditional univariate quality control frameworks. Conventional QC strategies often evaluate each assay independently, assuming that deviations in one variable do not influence others. However, molecular diagnostic assays frequently operate as multiplexed systems in which multiple viral targets are analyzed simultaneously. In such contexts, relationships among variables carry important information about system behavior. Multivariate statistical methods are therefore better suited to detect complex patterns of variability emerging from interacting analytical factors—a principle well established in pharmaceutical manufacturing under multivariate statistical process control (MSPC) theory [34,35], and now demonstrated here in a public health molecular diagnostic context.

Principal component analysis provides further evidence for the multidimensional nature of analytical variability in the dataset. The proportion of variance explained by the first principal component varied across viral targets, ranging from 54.1% in DENV4 to 100% in DENV3, indicating that different assays are governed by different combinations of variability sources. In some cases, a single dominant axis captured most of the variance, suggesting that analytical dispersion was driven by a common factor such as instrument calibration or reagent performance. In other cases, variance was distributed across multiple components, indicating the presence of several interacting factors. For ZIKV (PC2 = 34.1%) and CHIKV (PC2 = 28.8%), the substantial secondary component revealed systematic directional CT biases—consistent with findings from studies of RT-qPCR standard curve variability, which have shown that certain assay–platform combinations exhibit characteristic directional shifts independent of total efficiency [55].

This heterogeneity among assays suggests that diagnostic panels cannot always be treated as statistically equivalent. Even when assays share similar amplification protocols, their susceptibility to technical variation may differ depending on primer design, probe chemistry, target abundance and amplification efficiency. Studies examining RT-qPCR assay performance have shown that amplification efficiency differences can substantially affect quantitative interpretation, particularly when standard curves or calibration references are not consistently applied [24,53]. Consequently, surveillance systems that monitor multiple viral targets may benefit from assay-specific quality monitoring strategies rather than uniform QC thresholds.

The comparison between agglomerative (AGNES) and divisive (DIANA) hierarchical clustering also provides important methodological insight. Agglomerative clustering methods such as AGNES emphasize overall similarity structures within the data, gradually merging observations into larger groups, and are particularly useful for identifying stable analytical regimes. Divisive clustering methods such as DIANA begin with the entire dataset and recursively partition it by maximal internal dissimilarity, conferring greater sensitivity to early-stage deviations that have not yet become dominant features of the data structure [4,32]. The use of both clustering paradigms therefore allowed complementary perspectives on network behavior: AGNES identified stable clusters associated with instrument platforms and workflow configurations, while DIANA detected anomalous reagent lots that diverged from the dominant analytical structure before those deviations were visible in agglomerative analyses. The DENV-2 analysis exemplified this asymmetry: DIANA early-excised a suspect reagent lot in its first divisive step, treating it as maximally dissimilar from the core ensemble, whereas AGNES progressively incorporated the same lot into a higher-order cluster, effectively concealing its deviant character until later in the agglomeration hierarchy. This property has direct operational value for lot-qualification decisions prior to large-scale deployment of a suspect reagent batch.

The Baker’s gamma permutation test results (p = 0 for all six viral targets under N = 100 permutations) provide the statistical validation for this complementarity: both algorithms identify the same global cluster topology, even though they assign different internal leaf orderings within clusters. The extensive visual line-crossing observed in tanglegrams—often misinterpreted as evidence of algorithmic disagreement—in fact reflects this leaf-ordering divergence rather than topological conflict. This distinction is methodologically critical: a high tanglegram crossing density paired with a significant Baker’s gamma coefficient is the signature of two methods that independently confirm the same cluster structure while each revealing different aspects of within-cluster organization.

Another important dimension of quality surveillance concerns the detection of biologically implausible measurements. The dataset contained Ct values near zero (LACEN-AL) and approximately 18 cycles (LACEN-RO)—both outside the expected dynamic range of standardized RT-qPCR positive controls. Such measurements are unlikely to reflect true biological variation and instead indicate potential data integrity issues, including transcription errors, metadata misalignment or instrument misconfiguration. Similar anomalies have been reported in large-scale molecular diagnostic datasets, particularly during periods of rapid assay scale-up when information systems from multiple instruments are integrated under non-standardized data pipelines [56]. The presence of such values highlights the importance of incorporating automated data integrity validation into quality surveillance pipelines prior to multivariate analysis.

Process capability analysis provides additional insight into the temporal stability of laboratory performance. Capability indices capture both the dispersion of measurements and their alignment with predefined specification limits. In the present dataset, laboratories displayed heterogeneous capability metrics even when median Ct values appeared similar. Some laboratories exhibited tight control limits and highly predictable performance trajectories, whereas others showed broader variability or occasional drift events. This finding underscores a fundamental principle in quality science: process stability and process centering represent distinct dimensions of performance. A laboratory may produce measurements centered near expected values but exhibit substantial variability over time, reducing predictability and complicating corrective action. Conversely, a laboratory may display stable but systematically shifted measurements, indicating the need for recalibration or harmonization. Effective quality governance therefore requires simultaneous monitoring of both dimensions [42,43,44].

The broader implications of these findings extend beyond the specific context of arbovirus diagnostics. The COVID-19 pandemic demonstrated the central role of RT-qPCR in global infectious disease surveillance and highlighted the challenges associated with deploying molecular testing at unprecedented scale. Reviews of diagnostic testing during that period emphasize that interpreting RT-qPCR results requires careful consideration of both laboratory-based and real-world performance metrics, and that gaps in understanding fundamental diagnostic metrics have direct consequences for public health decision-making [56]. The experience of the LACEN network during the dengue epidemic illustrates similar dynamics under a different pathogen context, reinforcing that these are structural rather than disease-specific challenges for molecular diagnostic governance.

The network successfully maintained high testing throughput and sustained national arbovirus surveillance during a period of intense epidemiological pressure. This operational resilience represents a significant achievement for public health infrastructure. However, the presence of measurable analytical heterogeneity indicates that scaling diagnostic capacity does not automatically guarantee analytical harmonization. From a surveillance perspective, this distinction between capacity and harmonization is critical: diagnostic networks may produce large volumes of data during epidemics, but the epidemiological interpretation of those data depends on their comparability across laboratories. If Ct values are systematically shifted between laboratories, quantitative interpretations such as viral load estimation or threshold-based case classification may vary across jurisdictions, compromising the reliability of national epidemiological estimates [26,29].

These challenges highlight the need for more sophisticated quality surveillance frameworks in molecular diagnostic networks. Traditional QC systems, including Westgard rules [9], remain useful for identifying immediate run-level anomalies but operate primarily within a univariate paradigm. In contrast, modern diagnostic networks generate high-dimensional datasets containing multiple correlated variables, including Ct values across targets, instrument identifiers, reagent lots and operator metadata. Multivariate statistical methods are uniquely suited to extract information from such datasets. The analytical framework implemented in this study demonstrates how integrating clustering, discriminant analysis and principal component analysis can reveal structural relationships among laboratories, detect emerging anomalies and characterize the covariance structure of measurement variability—capabilities that remain invisible under conventional monitoring systems [47,48,49].

The use of standardized positive controls as the primary analytical substrate also warrants discussion. Positive controls provide a controlled environment for evaluating assay performance because they minimize biological variability and ensure that all laboratories analyze equivalent material. In this sense, they function as a diagnostic “reference signal” that reflects the technical backbone of the testing system. However, as detailed in section 5.1, positive controls cannot fully replicate the complexity of clinical samples, and future studies integrating clinical specimens with standardized controls may provide deeper insight into how analytical heterogeneity influences diagnostic sensitivity and specificity under real-world conditions.

Beyond the specific context of arboviral diagnostics, the findings presented here have broader implications for the governance of distributed molecular diagnostic systems globally. The COVID-19 pandemic highlighted the central role of RT-qPCR in infectious disease surveillance and exposed the challenges of maintaining analytical comparability across rapidly expanded diagnostic networks [56,57,58]. Reviews of diagnostic testing during that period emphasize that interpreting RT-qPCR results requires careful consideration of both laboratory-based and real-world performance metrics, and that gaps in understanding fundamental diagnostic metrics carry direct consequences for public health decision-making [56]. Similar dynamics occurred in the LACEN network during the dengue epidemic, reinforcing that these are structural rather than disease-specific challenges for molecular diagnostic governance. Distributed diagnostic systems require frameworks that conceptualize the network itself as a measurement ecosystem, and governing such ecosystems effectively demands analytical tools capable of operating at the network level [59,60,61].

The Brazilian LACEN network demonstrated remarkable operational resilience during the 2024 dengue epidemic, maintaining large-scale diagnostic throughput across geographically dispersed laboratories under conditions of extreme epidemiological pressure. This represents a significant public health achievement. However, the presence of measurable analytical heterogeneity underscores that scaling diagnostic capacity does not automatically ensure analytical harmonization. For epidemiological surveillance systems that integrate data from multiple laboratories, analytical harmonization is as important as analytical capacity. The multivariate QC framework presented here offers a practical pathway toward addressing this challenge: by integrating hierarchical clustering, discriminant analysis, principal component analysis, and process capability metrics within an automated pipeline, the system enables continuous monitoring of network-level diagnostic performance and could operate in near real time, identifying emerging analytical deviations before they influence surveillance outcomes [62,63].

In the context of climate change, urbanization, and expanding arboviral transmission zones, the demand for large-scale molecular diagnostic surveillance will likely continue to grow. The co-circulation of DENV1–4, ZIKV, and CHIKV observed in Brazil in 2024 exemplifies the complexity confronting national diagnostic networks as arboviral ecology shifts [11,27]. Strengthening the analytical governance of distributed laboratory networks therefore represents an increasingly important component of global health security. The analytical framework presented in this study demonstrates that advanced multivariate monitoring methods can be implemented using open-source statistical tools and applied successfully within real-world public health infrastructures—providing a replicable model for arboviral and broader infectious disease diagnostic QC governance during epidemic emergencies.

Despite these limitations, the findings presented here illustrate the value of integrating advanced statistical monitoring into public health diagnostic systems. The analytical pipeline developed for this study relies on widely available statistical tools and can be implemented within common computational environments such as R. With appropriate automation, such pipelines could operate in near real time, continuously analyzing incoming diagnostic data and alerting laboratory networks to emerging analytical deviations. In the context of epidemic preparedness, such capabilities may prove increasingly important: as emerging infectious diseases continue to require rapid expansion of diagnostic capacity across geographically distributed laboratories, ensuring analytical comparability becomes essential for accurate epidemiological modeling, resource allocation and public health decision-making.

In conclusion, the 2024 dengue epidemic provided a unique opportunity to examine the behavior of a national molecular diagnostic network operating under extreme epidemiological pressure. By applying a comprehensive multivariate surveillance framework to positive-control CT data from 15 LACENs, this study demonstrates that while the Brazilian network maintained strong operational capacity and internal laboratory stability, measurable analytical heterogeneity persisted across laboratories at the network level. Conceptualizing distributed diagnostic networks as measurement ecosystems—in which CT values emerge from the interaction of instruments, reagents, operators, and algorithms—provides a theoretically coherent framework for understanding and addressing this heterogeneity. These findings emphasize the importance of distinguishing between diagnostic throughput and analytical harmonization, and highlight the potential of multivariate statistical surveillance as a practical, open-source-implementable tool for strengthening molecular diagnostic governance in large public health infrastructures facing epidemic pressures.

## Limitations

### 5.1 Scope: Positive Controls vs. Clinical Specimens

The most fundamental scope limitation of this study is that all analyses were conducted exclusively on positive-control CT data. Positive controls are standardized RNA preparations of defined concentration used to verify assay function, and their CT behavior provides a necessary but not sufficient indicator of performance on actual clinical specimens. Clinical specimens introduce additional analytical challenges absent from positive controls: endogenous PCR inhibitors co-extracted with viral RNA, variability in RNA degradation rates across specimen types and storage conditions, competitive amplification from host RNA, and matrix-specific extraction efficiency differences [24,29].

The relationship between positive-control CT performance and clinical specimen analytical accuracy is not guaranteed to be linear or universal across all failure modes. A laboratory with stable positive-control CTs may still underperform on clinical specimens if inhibitor removal is suboptimal or if extraction efficiency is systematically low. Conversely, a laboratory with moderately elevated positive-control CTs (consistent with a calibration offset) may perform acceptably on clinical specimens if the offset is consistent and reproducible. Positive-control surveillance is therefore best understood as an early-warning indicator for assay integrity rather than a direct measurement of diagnostic accuracy on patient samples.

### 5.2 Classification Accuracy and Overlapping Laboratory Profiles

Cross-validated LDA and QDA classification accuracies of 37.7% and 49.1%, respectively, fall substantially short of levels required for reliable standalone laboratory identification. While both values significantly exceed chance expectation, the remaining misclassification rate reflects genuinely overlapping multivariate CT profiles among laboratories that operate similar platforms and reagent lots—which is, analytically speaking, the desirable state for a well-harmonized network. The discriminant approach therefore identifies analytical divergence rather than measuring network performance, and its classification ceiling should be interpreted accordingly.

Future studies incorporating richer metadata covariates—RNA extraction platform manufacturer and lot, ambient temperature at time of assay, operator certification level and years of experience, thermocycler age and most recent calibration date—would likely improve classification accuracy while simultaneously providing mechanistically informative loadings on the discriminant axes. A mixed-effects model framework incorporating these covariates as fixed effects, with laboratory as a random effect, would complement the current discriminant approach by formally partitioning CT variance across known and unknown sources.

### 5.3 Incomplete Coverage Across Viral Targets and Statistical Tests

The formal statistical testing framework—ANOVA, Levene’s test, Shapiro–Wilk, and regression analysis—was fully presented only for ZIKV × operator in this study. Equivalent analyses for CHIKV, DENV1, DENV2, DENV3, and DENV4 remain to be completed in subsequent pipeline iterations. The asymmetry in coverage limits the generalizability of the operator-effect conclusions to ZIKV specifically and warrants systematic extension across all viral targets.

Additionally, the non-normality of residuals (Shapiro–Wilk p ≈ 0) and heteroskedasticity (Levene p ≈ 0) indicate that the parametric ANOVA results for ZIKV should be validated with complementary non-parametric tests. Kruskal–Wallis testing and Welch ANOVA (robust to unequal variances) are the recommended alternatives and should be incorporated into all future pipeline runs. Games–Howell post-hoc testing (appropriate under heteroskedasticity) is recommended in place of Tukey’s HSD for pairwise operator comparisons.

### 5.4 Absence of Pre-Epidemic Baseline

The study period did not include a formal pre-epidemic quality baseline for comparison. Without inter-epidemic reference data, it is not possible to formally test whether quality performance deteriorated systematically during the epidemic surge—a question of direct relevance for epidemic preparedness planning. Longitudinal QC data from inter-epidemic periods would enable formal changepoint analysis to identify when quality metrics first began deviating from baseline, whether that deviation recovered as the epidemic declined, and whether different failure modes were temporally associated with different phases of the outbreak cycle.

Future iterations of this surveillance framework should prioritize prospective data collection extending into inter-epidemic periods, enabling formal before–after comparisons and the development of epidemic-stress-adjusted quality tolerance limits.

### 5.5 Geographic Analysis Not Executed

The bioinformatic pipeline included code for geographic analysis stratified by state (Acre, Amazonas, Paraíba, Piauí, Pernambuco, and others), but this analysis was not executed in the current iteration. Geographic analysis would enable detection of regional clustering patterns in CT performance—potentially revealing whether analytical quality correlates with geographic features such as climate zone (affecting reagent stability), infrastructure tier (affecting equipment maintenance), or distance from centralized training centers (affecting operator skill). This analytical layer is technically feasible within the existing pipeline and represents a high-priority next step for subsequent QC surveillance rounds.

### 5.6 Non-parametric Validation Pending

As noted above, the parametric analytical framework (ANOVA, LDA, regression) was applied to data confirmed to violate normality and homoskedasticity assumptions. While the large sample size (n = 3,192) provides some robustness to these violations under the central limit theorem, the degree of non-normality (W = 0.85) is substantial enough that parametric p-values may be liberal. A comprehensive non-parametric validation—including permutation-based ANOVA, bootstrap confidence intervals for regression coefficients, and rank-based concordance measures—would strengthen the inferential claims of this study and is recommended as a standard component of future pipeline runs.

### 5.7 Reagent Lot Qualification Not Prospective

The identification of anomalous reagent lots through this analysis was conducted retrospectively on data already used in clinical testing. While retrospective identification is analytically valuable for root-cause analysis and future lot procurement decisions, it does not protect patients tested with the deviant lots during the detection window. A prospective implementation of the DIANA-based lot qualification strategy—where incoming lots are characterized by a pilot panel of positive-control runs before network-wide deployment—would transform the current retrospective surveillance tool into a genuine preventive quality mechanism. The feasibility of such prospective qualification within the LACEN network’s operational constraints deserves formal evaluation in a follow-up implementation study.

## Conclusions

This multicenter QC study, conducted across 15 Brazilian LACENs during the unprecedented 2024 dengue epidemic, provides comprehensive empirical evidence for the analytical performance and sources of variability in RT-qPCR molecular diagnostics for DENV1–4, ZIKV, and CHIKV. Analysis of 3,192 complete RT-qPCR runs and 19,152 datapoints through an integrated multivariate statistical framework revealed a diagnostic network characterized by locally stable but globally heterogeneous analytical processes.

Five principal conclusions emerge from this work.

1. PCR instrument platform and reagent lot identity, not geographic location or laboratory size, are the dominant structural determinants of inter-laboratory CT variability across the LACEN network. This finding is consistent across hierarchical clustering, PCA, and heatmap analyses. Quality governance interventions should therefore prioritize inter-platform CT harmonization protocols and shared lot-qualification procedures over geographic or administrative reorganization.
2. The integrated multivariate framework—encompassing Baker’s gamma permutation testing, PVClust bootstrap analysis, PCA, LDA/QDA, and process capability metrics—substantially outperforms univariate Westgard-rule monitoring in detecting emergent assay drift, identifying outlier lots and operators, and providing mechanistic hypotheses about quality failure sources. The two approaches are complementary, not competing, and their integration into a layered quality architecture provides the most comprehensive analytical protection.
3. Operator-specific CT offsets of up to +4.0 cycles carry direct clinical consequences for borderline specimen classification. ANOVA and regression analysis confirmed highly significant operator effects on ZIKV CT values (F = 8.799, p ≈ 0), with identifiable operator-specific deviations representing actionable targets for evidence-based retraining. Equivalent analyses across all viral targets are warranted and represent a priority for future pipeline iterations.
4. The identification of biologically implausible extreme CT values (CT ≈ 0; CT ≈ 18) underscores the need for automated pre-analysis range-checking and a formal data integrity protocol as prerequisites for valid QC assessment. These anomalies, if undetected, distort all downstream multivariate analyses and may generate false epidemiological signals.
5. The successful deployment of a fully automated R-based bioinformatic pipeline for multivariate QC across a geographically dispersed, low-resource laboratory network demonstrates operational feasibility under real-world epidemic conditions. As arboviral diseases continue to expand their geographic range under climate change and urbanization pressures, the lessons drawn from this study—the primacy of instrument harmonization, the complementarity of univariate and multivariate QC, the clinical significance of operator variability, and the value of automated bioinformatic quality governance—are of immediate and growing global relevance.

## Supporting Information

S1 Dataset. Compiled positive-control CT dataset across 15 LACENs, 2024 epidemic year. Tab-delimited file including run ID, date, operator identifier, reagent lot, thermocycler model, laboratory code, and CT values for DENV1–4, ZIKV, and CHIKV. Personal identifiers have been removed. Available from the corresponding author upon reasonable request pending data sharing agreement with the Brazilian Ministry of Health.

S2 Code. Annotated R bioinformatic pipeline for all analyses described in the Methods section. Includes scripts for data preprocessing, AGNES/DIANA clustering, Baker’s gamma permutation testing, PVClust, PCA, LDA/QDA, ANOVA, CEP charts, process capability analysis, and ComplexHeatmap visualization. Available as a GitHub repository (link to be provided upon acceptance).

## Data Availability Statement

The data underlying this study are positive-control RT-qPCR records generated by the Brazilian Central Public Health Laboratory (LACEN) network in the context of routine epidemic surveillance. Primary data access is governed by the Brazilian Ministry of Health (Coordenação-Geral de Laboratórios de Saúde Pública — CGLAB/SVS) and is subject to institutional data governance agreements. Researchers interested in accessing the dataset should contact the corresponding author and the relevant LACEN institutional representatives. The full analytical R pipeline (S2 Code) will be made publicly available on GitHub upon acceptance of the manuscript.

## Author Contributions

Conceptualization: Emerson Luiz Lima Araújo, Daniel Ferreira de Lima Neto.

Data curation: Ludmila Oliveira Carvalho Sena, Jayra Juliana Paiva Alves Abrantes, Mariana Araújo Costa, and all co-authors at participating LACENs.

Formal analysis: Daniel Ferreira de Lima Neto.

Investigation: Ludmila Oliveira Carvalho Sena, Jayra Juliana Paiva Alves Abrantes, Mariana Araújo Costa, and all participating LACEN teams.

Methodology: Daniel Ferreira de Lima Neto.

Project administration: Emerson Luiz Lima Araújo, Daniel Ferreira de Lima Neto. Supervision: Emerson Luiz Lima Araújo, Daniel Ferreira de Lima Neto.

Visualization: Daniel Ferreira de Lima Neto.

Writing – original draft: Emerson Luiz Lima Araújo, Daniel Ferreira de Lima Neto.

Writing – review & editing: Ludmila Oliveira Carvalho Sena, Jayra Juliana Paiva Alves Abrantes, Mariana Araújo Costa, and all co-authors.

## References

[1] Lun ATL, McCarthy DJ, Marioni JC. (2019). A step-by-step workflow for low-level analysis of single-cell RNA sequencing data with Bioconductor. F1000Research, 5, 2122.

[2] Westgard JO. (2010). Basic QC Practices. 3rd ed. Madison, WI: Westgard QC Inc.

[3] CAMO Analytics. (2019). Multivariate Data Analysis and Design of Experiments for Biotechnology. CAMO Software White Paper.

[4] Kaufman L, Rousseeuw PJ. (2009). Finding Groups in Data: An Introduction to Cluster Analysis. Wiley-Interscience, New York.

[5] Alon U, Barkai N, Notterman DA, et al. (1999). Broad patterns of gene expression revealed by clustering analysis of tumor and normal colon tissues. PNAS, 96(12), 6745–6750.

[6] Li Y, Ge X, Peng F, Li W, Li JJ. (2022). Exaggerated false positives by popular differential expression methods when analyzing human population samples. Genome Biology, 23, 79.

[7] Tibshirani R, Hastie T, Narasimhan B, Chu G. (2002). Diagnosis of multiple cancer types by shrunken centroids of gene expression. PNAS, 99(10), 6567–6572.

[8] Dudoit S, Fridlyand J, Speed TP. (2002). Comparison of discrimination methods for the classification of tumors using gene expression data. JASA, 97(457), 77–87.

[9] Westgard JO, Barry PL, Hunt MR, Groth T. (1981). A multi-rule Shewhart chart for quality control in clinical chemistry. Clinical Chemistry, 27(3), 493–501.

[10] Gurgel-Gonçalves R, de Oliveira WK, Croda J. (2024). The greatest dengue epidemic in Brazil. Rev Soc Bras Med Trop, 57, e00203–2024.

[11] Daude MM, Manuli ER, Pereira GM, et al. (2024). Simultaneous detection of arboviruses by a multiplex RT-qPCR assay in Tocantins. Braz J Infect Dis, 28(4), 103855.

[12] Wickham H, et al. (2023). dplyr: A Grammar of Data Manipulation. R package version 1.1.4.

[13] Maechler M, et al. (2022). cluster: Cluster Analysis Basics and Extensions. R package version 2.1.4.

[14] Kassambara A, Mundt F. (2020). factoextra: Extract and Visualize the Results of Multivariate Data Analyses. R package version 1.0.7.

[15] Venables WN, Ripley BD. (2002). Modern Applied Statistics with S. 4th ed. Springer, New York.

[16] Fox J, Weisberg S. (2019). An R Companion to Applied Regression. 3rd ed. SAGE Publications.

[17] Gross J, Ligges U. (2015). nortest: Tests for Normality. R package version 1.0-4.

[18] Scrucca L. (2004). qcc: An R package for quality control charting. R News, 4(1), 11–17.

[19] Tang Y. (2020). ggQC: Quality Control Charts for ggplot. R package version 0.0.31.

[20] Wickham H. (2016). ggplot2: Elegant Graphics for Data Analysis. Springer-Verlag, New York.

[21] Zhu H. (2021). kableExtra: Construct Complex Table with kable and Pipe Syntax. R package version 1.3.4.

[22] Grolemund G, Wickham H. (2011). Dates and times made easy with lubridate. Journal of Statistical Software, 40(3), 1–25.

[23] Gentle JE. (2007). Matrix Algebra: Theory, Computations, and Applications in Statistics. Springer, New York.

[24] Bustin SA, Benes V, Garson JA, et al. (2009). The MIQE guidelines. Clinical Chemistry, 55(4), 611–622.

[25] Sena BF, Herrera BB, et al. (2024). Performance of CDC Trioplex qPCR during a dengue outbreak in Brazil. Braz J Infect Dis, 28(3), 103766.

[26] Barra G, Vieira A, Lima S, et al. (2024). Evaluating RT-qPCR for Dengue Virus Detection: Outcomes of a Brazilian External Quality Assessment Program. Clinical Chemistry, 70, hvae106.575.

[27] Mishra N, Ng J, Rakeman JL, et al. (2019). One-step pentaplex real-time PCR assay for detection of Zika, dengue, chikungunya, West Nile viruses. J Clin Virol, 120, 44–50.

[28] Versiani AF, et al. (2023). Performance of VIDAS Diagnostic Tests for Dengue NS1 Antigen and Anti-Dengue IgM/IgG. Diagnostics, 13(6), 1137.

[29] Livak KJ, Schmittgen TD. (2001). Analysis of relative gene expression data using real-time quantitative PCR and the 2-DDCT method. Methods, 25(4), 402–408.

[30] Nolan T, Hands RE, Bustin SA. (2006). Quantification of mRNA using real-time RT-PCR. Nature Protocols, 1(3), 1559–1582.

[31] Huggett JF, Foy CA, Benes V, et al. (2013). The digital MIQE guidelines. Clinical Chemistry, 59(6), 892–902.

[32] Milligan GW, Cooper MC. (1987). Methodology review: Clustering methods. Applied Psychological Measurement, 11(4), 329–354.

[33] Suzuki R, Shimodaira H. (2006). Pvclust: an R package for assessing the uncertainty in hierarchical clustering. Bioinformatics, 22(12), 1540–1542.

[34] Kourti T, MacGregor JF. (1995). Process analysis, monitoring and diagnosis, using multivariate projection methods. Chemometrics Intell Lab Syst, 28(1), 3–21.

[35] Ahsan M, et al. (2022). Kernel PCA control chart for monitoring mixed non-linear variable and attribute quality characteristics. Sci Rep, 12, 15723.

[36] ANVISA. (2005). RDC n° 302. Regulamento Técnico para funcionamento de Laboratórios Clínicos. Brasília.

[37] Burdick RK, Borror CM, Montgomery DC. (2003). A review of methods for measurement systems capability analysis. J Qual Technol, 35(4), 342–354.

[38] Venkatraman V, Kramer N. (2010). Multivariate quality control in the clinical laboratory: The role of LDA and PCA. Clinical Biochemistry, 43(4–5), 432–439.

[39] AIAG. (2002). Measurement Systems Analysis Reference Manual. 3rd ed. Automotive Industry Action Group.

[40] Huber PJ. (1981). Robust Statistics. Wiley, New York.

[41] Rousseeuw PJ, Leroy AM. (1987). Robust Regression and Outlier Detection. Wiley, New York.

[42] Montgomery DC. (2020). Introduction to Statistical Quality Control. 8th ed. Wiley, New York.

[43] Taylor S, et al. (2010). A practical approach to RT-qPCR—publishing data that conform to the MIQE guidelines. Methods, 50(4), S1–S5.

[44] Westgard JO. (2023). Westgard Sigma Rules. Westgard QC. https://westgard.com

[45] Goswami B, et al. (2022). Sigma metrics as a tool for evaluating internal quality control in clinical chemistry. Indian J Clin Biochem, 37, 1–7.

[46] Wang Y, Li X, Li D, et al. (2024). A modified quality control protocol for infectious disease serology based on the Westgard rules. Sci Rep, 14, 16683.

[47] Mahalanobis PC. (1936). On the generalized distance in statistics. Proc Natl Inst Sci India, 2(1), 49–55.

[48] Jackson JE. (2003). A User’s Guide to Principal Components. Wiley, New York.

[49] Shinto M, Kaji T, Asano K. (2005). Multivariate statistical process control in clinical laboratories using the Mahalanobis-Taguchi system. J Clin Lab Anal, 19(1), 1–8.

[50] Pires de Miranda M, et al. (2010). 2nd International External Quality Control Assessment for Molecular Diagnosis of Dengue. PLOS Negl Trop Dis, 4(10), e833.

[51] Eisen MB, Spellman PT, Brown PO, Botstein D. (1998). Cluster analysis and display of genome-wide expression patterns. PNAS, 95(25), 14863–14868.

[52] Dellière S, Gits-Muselli M, White PL, et al. (2020). Quantification of Pneumocystis jirovecii: Cross-Platform Comparison of One qPCR Assay with Leading Platforms and Six Master Mixes. J Fungi (Basel), 6(1), 9. 10.3390/jof6010009

[53] Bilgrau AE, Falgreen S, Petersen A, et al. (2016). Unaccounted uncertainty from qPCR efficiency estimates entails uncontrolled false positive rates. BMC Bioinformatics, 17, 159. 10.1186/s12859-016-0997-6

[54] Pfaffl MW. (2001). A new mathematical model for relative quantification in real-time RT-PCR. Nucleic Acids Research, 29(9), e45. 10.1093/nar/29.9.e45

[55] De Oliveira GH, et al. (2025). The Impact of the Variability of RT-qPCR Standard Curves on Reliable Viral Detection in Wastewater Surveillance. Microorganisms, 13(4), 776. 10.3390/microorganisms13040776

[56] Bustin SA. (2024). RT-qPCR Testing and Performance Metrics in the COVID-19 Era. International Journal of Molecular Sciences, 25(17), 9326. 10.3390/ijms25179326

[57] Peeling RW, Olliaro PL, Boehme CC, et al. (2020). Diagnostics in epidemic response. Lancet Infectious Diseases, 20(9), e199–e208. 10.1016/S1473-3099(20)30274-4

[58] Mina MJ, Parker R, Larremore DB. (2020). Rethinking Covid-19 Test Sensitivity — A Strategy for Containment. New England Journal of Medicine, 383(22), e120. 10.1056/NEJMp2025631

[59] Pai M, Ghoshal S. (2015). Diagnostic networks for global health. PLOS Medicine, 12(9), e1001878. 10.1371/journal.pmed.1001878

[60] Drain PK, Ampajwala M, Chaudhri C, et al. (2019). A need for point-of-care diagnostics for infectious diseases in resource-limited settings. Nature Reviews Microbiology, 17(7), 415–424. 10.1038/s41579-019-0192-y

[61] Vandenberg O, Martiny D, Rochas O, van Belkum A, Kozlakidis Z. (2021). Considerations for diagnostic COVID-19 tests. Nature Reviews Microbiology, 19(3), 171–183. 10.1038/s41579-020-00461-z

[62] Mishra N, Ng’eno EC, Bhatt KN, et al. (2019). Multiplex PCR-based diagnosis of arboviral infections. Journal of Clinical Virology, 121, 104212. 10.1016/j.jcv.2019.104212

[63] Versiani AF, Codeço CT, Coelho FC, et al. (2023). Dengue diagnostic performance across platforms: implications for surveillance. Diagnostics (Basel), 13(4), 649. 10.3390/diagnostics13040649

